# Design of a Clinical Balance Tool for Fall Risk Assessments: A Development and Usability Study

**DOI:** 10.1101/2024.03.28.24305053

**Authors:** Jennifer Hornung Garvin, Virginia M. Yazzie, Natalie A. Katsuyama, Truman Rudloff, Lise C. Worthen-Chaudhari, Ajit M.W. Chaudhari

## Abstract

**Background:** Falls are a significant source of early morbidity and mortality in the aging population, yet clinical changes that lead to increased fall risks often escape early identification and intervention. A device to measure postural control would facilitate evidence-based fall risk assessment.

**Objectives:** Our objectives were to iteratively develop a prototype quantitative posture instrument (QuPI) to replace the weight scale and to assess barriers and facilitators of its implementation in a clinical setting.

**Methods:** We undertook a formative evaluation and usability study of two QuPI prototypes in primary care, medical oncology, sports medicine, cardiology, and endocrinology outpatient clinics. Clinicians evaluated an initial QuPI prototype and completed a semi-structured interview to determine critical functionality, inform design, and assess usability. The QuPI was modified according to the results, and a new prototype was tested and evaluated.

**Results:** Eighteen clinicians participated in both rounds of interviews. Clinicians who participated (referred to as participants) reported willingness to use the QuPI with all patients during the first round of interviews and stated they would replace their current weight scale with the modified QuPI during the second round of interviews. Participants identified design elements that were both facilitators and barriers to use. Usability scores for both prototypes were excellent. Despite several national guidelines for fall risk assessments, lack of consistent use of guidelines by care teams was found to be a barrier to effective fall risk assessments.

**Conclusion:** The QuPI provides a new method for quantifying fall risks with good user acceptance, usability, and clinical feasibility without disrupting workflow. The QuPI supplemented and facilitated the use of standard algorithms for fall risk assessment. Greater education of the entire care team regarding evidence-based fall risk assessment will promote adherence to guidelines and fall prevention.

## Introduction

Falls are a significant source of early morbidity and mortality in the aging population[1], yet the neurological, sensory, and motor changes that lead to increased fall risks often escape early identification and intervention. Vital signs are commonly used in clinical settings to assess the cardiovascular system (blood pressure, heart rate), immune system (body temperature), and respiratory system (respiratory rate) to establish baseline values, screen for increased risk of co-morbidities or disease, and enable changes from baseline to be identified and communicated across times and locations[2]. There is no simple vital sign that can be used to assess the balance system, which draws upon neurological, sensory, and motor functions. Although clinical measurement of balance can predict the risk of falls[3–6], clinical balance testing is not common practice in primary care settings because of time constraints, lack of awareness, and other factors.

The prevalence and consequences of injurious falls present an urgent need to capture data for use as a vital sign for the balance system that is inexpensive, easy to adopt, consistent, and objective, with an evidence base supporting its sensitivity and specificity. Quantitative posture control is a good candidate for such a vital sign. Quantitative measurement of posture control is faster, more sensitive, and more specific than clinical balance testing for fall risk assessment[7–9]; however, both approaches face barriers to adoption in clinical practice, including time requirements, expense, lack of awareness, and need for significant technical expertise.

The purposes of this qualitative interview-based study were to 1) characterize the workflow of patient intake, vital sign assessment, and fall risk assessment in physician or advanced practice provider clinics and 2) identify design requirements for a quantitative posture instrument (QuPI) to efficiently measure postural control in these settings.

## Methods

### Theoretic Frameworks

To develop a QuPI as a Health Information Technology (HIT) tool for clinical practice, we undertook a formative evaluation and usability assessment using three frameworks: the HIT sociotechnical framework[10], the Promoting Action on Research Implementation in Health Services (PARIHS) framework[11], and the Cabana framework[12]. The HIT sociotechnical framework[10] emphasizes characteristics of the clinical and organizational contexts in which the tool will be implemented. The PARIHS framework considers barriers and facilitators to the intended use of the tool[11]. The Cabana framework identifies provider-related dimensions to explain barriers (e.g., gaps in knowledge, attitudes, or practice) related to adherence to clinical practice guidelines[12]. Together, these frameworks informed the design of the QuPI in terms of understanding the human, clinical, and overall sociotechnical contexts in which the QuPI will be used.

### Study Design

This was a descriptive formative evaluation study. Clinicians who participated (referred to as participants) evaluated an initial prototype of the QuPI in a semi-structured interview. A modified QuPI prototype was then created, and the participants evaluated the modified prototype in a second semi-structured interview. The QuPI prototypes were used with clinical scenarios for formative evaluation and usability assessment (**Fig 1 The organizational flow of the study**).

**Figure 1.**
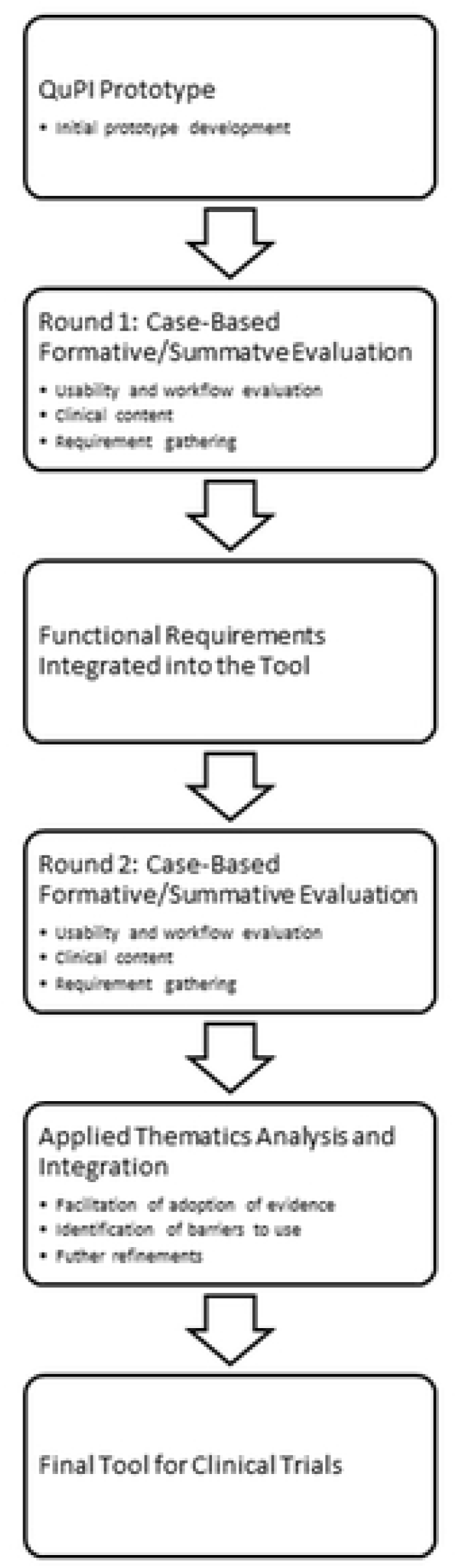

### Settings and Description of the Initial QuPI Prototype

The research was conducted in outpatient clinics including primary care, cardiology, oncology, endocrinology, and sports medicine practices within an academic healthcare organization and a large primary care network. The QuPI initial prototype was based on a standard balance platform similar to those commonly used in studies of postural sway[13–16], except that its dimensions were chosen to match the length and width of the weight scales used in most clinics (40 cm × 33 cm), but with a lower height (4.4 cm). The user interface of the initial prototype indicated the weight of the patient and had an onscreen button to start a 60-second timer for balance measurement. Upon completion, the display gave a number representing the balance score, which was calculated as the root-mean-square deviation of the center of pressure in the medial-lateral direction [7,13,14,17]. The device was accompanied by an information card with instructions to be read to the patient and a visual scale to interpret the balance score as low risk, medium risk, high risk, or immediate risk of a fall. The modified version of QuPI used in the second round of interviews had the following additional design features: reduced time required for the test from 60 seconds to 30 seconds; extended time for the score and weight display on the screen; an audible tone when timer expires; and display of balance score with tenths, not just the integer value. The instruction card was also modified to include notation that it is permissible to tell the patient how much time is left (halfway done, 15 seconds to go, 10 seconds to go, etc.) and that they are doing a good job.

### Human Subject Protection

This study was approved by The Ohio State University Biomedical IRB. The initial approval was provided on October 30, 2018 with the start of prospective recruiting on November 8, 2018 and the end of recruitment June 30, 2019. Informed consent was obtained verbally with a documentation of consent waiver approved by the IRB. There were no minors participating in this study.

### Participants

We sought a diverse group of clinicians as participants from academic and non-academic medical centers. The participants were not part of the study team and came from primary care, sports medicine, and other specialty practices in two healthcare systems. In this study, “providers” are defined as physicians and nurse practitioners, and “support staff” are defined as medical assistants and physician extenders.

### Procedure

We conducted two rounds of semi-structured interviews in the participants’ practice settings (**Table 1**).

**Table 1.** Evaluation questions used to guide assessment of the QuPI.

| Evaluation Component | Evaluation Questions |
| --- | --- |
| Guideline-directed care | Was the clinical content relevant? |
|  | Describe your knowledge of the guidelines. |
|  | Describe barriers to the use of the guidelines. |
| Support | Was the tool aligned with workflow? |
| Adoption and uptake of evidence using Health Information Technology | Did the tool increase awareness of the indications of falls risk and facilitate the use of treatment evidence as expressed in the ordering? |
| End-user design effectiveness | Was the QuPI useful? |
| Usability | Was the human interface with the QuPI well designed? |

The participants were able to use an initial prototype of the QuPI during the first interview and then a modified prototype during the second interview. These prototypes were used with clinical scenarios for formative evaluation and usability assessment. We also asked the participants to describe and demonstrate their work process for collecting vital signs from patients. Responses were captured via written notes and video/audio recordings. Audio recordings of the participants’ responses were transcribed and used to conduct an applied thematic analysis in which two researchers independently reviewed the transcripts, identified snippets to inform redesign, iterated codes in three to four cycles to evaluate concepts from the theoretical framework, identified thematic domains, and developed themes [18–19]. The whole research team then reviewed the themes (illustrated by snippets), refined them, and provided them to the design team for modification of the QuPI.

We used the System Usability Scale (SUS) [20–22] during the interviews to assess the usability of the QuPI. The SUS was composed of 10 questions scored on a five-point scale from ‘strongly agree’ to ‘strongly disagree.’ The final overall scaled score of the SUS had a possible range from 0 to 100, where higher scores indicate greater usability of the QuPI by end-users (**Table S1**). The grading analog for the SUS is supported by an adjective rating scale and quartile and acceptability ranges [22]. A score of 80.3 or higher was considered an ‘A’ (top 10% of scores), allowing users to recommend the QuPI to others [22]. The interview and SUS results were used to iteratively redesign the QuPI between the first and second rounds of interviews (**Fig 1**).

### Interviews

The semi-structured interviews were developed using theoretical models [10,11,12] (**Table 2**).

**Table 2.** Theoretical models used in this study and the aspects in which they were applied.

| Theoretical Model | Application Areas |
| --- | --- |
| Promoting Action on Research in Implementation of Health Services [11] | • Evidence-based research<br>• Context of implementation<br>• Facilitation techniques to overcome known barriers to implementation |
| Sociotechnical Model for Health Information Technology [10] | • Hardware and software<br>• Workflow and communication<br>• Clinical content<br>• Human-computer interface (usability) |
| Cabana Framework [12] | • Providers' attitudes, knowledge, and behaviors regarding clinical evidence |

Information concerning the participants’ knowledge of fall risk assessments and their current ability to determine fall risk was gathered to identify requirements and assess workflow to iteratively develop the QuPI prototype. The initial and modified QuPI prototypes were used with clinical scenarios for formative evaluation and usability assessment. We explored the usability of the QuPI in clinical settings [20–22] and identified features needed by end-users to incorporate the QuPI into routine clinical practice and clinical decision-making. We also explored the alignment of the QuPI with current processes for weight-measurement devices and explored barriers to the use of the QuPI in terms of hardware and software for electronic health records (EHR) systems, workflow and communication of the care team, and clinical guidelines [10,11].

## Results

### Participants

A total of 18 participants (5 male, 13 female) completed the first and second rounds of interviews and assessments. The participants included three primary care physicians, three physicians with other specialty practices, two sports medicine physicians, one primary care nurse practitioner, one nurse practitioner with a different specialty practice, two primary care medical assistants, two medical assistants with a different specialty practice, and four sports medicine physician extenders.

### Fall Risk Assessment and Documentation

We evaluated the workflow as described in the HIT sociotechnical framework [10] to determine the current state of fall risk assessments in the clinics and inform the development of the QuPI. The participants assessed fall risks by a variety of methods at several locations within their clinical practice. Fall risks were sometimes documented in the medical record as well as outside the medical record via notes, verbal information, and behavioral cues. In the first round of interviews, participants indicated that they assessed fall risks via pre-visit review of medications (11%, 2/18), patient reports (44%, 8/18), review of the vital signs flowsheet (11%, 2/18), observation by a member of the care team before and during the examination (83%, 15/18), clinician questioning (77%, 14/18), and review of the EHR template (11%, 2/18). The instruments used to assess fall risks included clinical guidelines (22%, 4/18), observation (72%, 13/18), a fall risk scale (5%, 1/18), a fall risk calculator (22%, 4/18), ad hoc questioning (44%, 8/18), and tests for specific underlying conditions that affect balance (33%, 6/18), including auscultation, electrocardiography, blood pressure measurement for cardiovascular conditions, deep tendon reflex and monofilament tactile sensation assessment for neurosensory deficits, and static visual acuity assessment for visual deficits.

The results of fall risk assessments were documented in several locations, including medical assistant notes (77%, 14/18), the flowsheet or vital signs package of the EHR (72%, 13/18), structured questions about fall risk within the medical record or on paper (16%, 3/18), and free-text notes within a set of structured data related to fall risk (16%, 3/18). Fall risk was also noted using an EHR template (22%, 4/18), in the chief complaint (50%, 9/18), in the History and Physical Exam (16%, 3/18), or in progress notes or other provider documents (61%, 11/18). Other communication occurred via verbal or written information given to providers by other team members outside of the medical record (16%, 3/18). This communication from the team was sometimes (22%, 4/18) incorporated later into the provider notes within the EHR.

### Knowledge of and Adherence to Clinical Guidelines

We assessed the use of clinical guidelines as part of the HIT sociotechnical framework[10] to inform the use of the QuPI. Participants expressed agreement with existing clinical guidelines but also described limitations related to the guidelines. They reported general knowledge of evidence-based fall risk assessments but often lacked detailed knowledge of the guidelines (**Table S2** and **Table S3**). The participants often prioritized other injuries or concerns over fall risks because of a lack of time or because the primary purpose of the visit was something other than concern about fall risk.

A majority (67%, 12/18) of the participants knew or had heard of at least one or more of the nationally promoted algorithms from the Centers for Disease Control (STEADI), the Johns Hopkins Healthcare System (JHFRAT), or the American Geriatric Society (AGS) [23–25]; however, none (0/6) of the subset of participants specializing in sports medicine had heard of any of these algorithms. Only two of the participants reported having undergone previous specialized training to work with geriatric patients.

### Recommendations for Modification of the QuPI and its Use in Clinical Practice

In the first round of interviews, the majority (89%, 16/18) of participants suggested features that could be added to the initial QuPI prototype and provided recommendations. Time constraints, the size of the device, the size of the information displayed, and difficulty using the device with patients who were obese and/or had ambulation difficulties were reported as potential barriers to routine use of the initial QuPI prototype. The low height of the device, ease of use and interpretation of results, and use of the device for both weight and balance measurement were reported as potential facilitators of routine use (**Table S4**).

In the second round of interviews, the participants expressed broad acceptance of the modified device and said they would use the device with all their patients in place of a conventional weight scale (**Table S4**). The changes made to the initial QuPI prototype are described in the settings section above based on the feedback described below. Participants suggested in the second round of interviews that it would be easier for patients to stand still for the measurement if verbal feedback about how much time was remaining was given every 10 to 15 seconds. Several participants also suggested the duration of the measurement should be shortened further if possible, and that there should be handrails placed around the device to make it safer to use by patients with balance problems. Another suggestion for further improvement was that the values shown on the screen could be given with green-yellow-red color coding to make it easier to quickly interpret the results.

### SUS Reports

Two SUS reports were conducted for the initial and final designs of QuPI. The initial mean SUS score was 83.57. The final QuPI prototype achieved a mean SUS score of 86.80, indicating a system with excellent usability (**Table 3**).

**Table 3.** System Usability Scale [20–22] results from the first and second interviews.

| Summative/Formative Evaluation Questions | 1<sup>st</sup> Interview<br>(average, n=18) | 2<sup>nd</sup> Interview<br>(average, n=18) |
| --- | --- | --- |
| 1. I would like to use the QuPI. | 4 | 5 |
| 2. I find the QuPI to be unnecessarily complex. | 2 | 1 |
| 3. I think the QuPI is easy to use. | 5 | 5 |
| 4. I would need the support of a technical person to be able to use the QuPI. | 1 | 1 |
| 5. I think the QuPI functions are well integrated. | 4 | 4 |
| 6. I think there is too much inconsistency in the QuPI. | 2 | 4 |
| 7. I imagine that most people would get used to the QuPI very quickly. | 5 | 5 |
| 8. I find the QuPI to be cumbersome to use. | 2 | 1 |
| 9. I am confident using the QuPI. | 5 | 4 |
| 10. I needed to know a lot more about the QuPI before I felt comfortable using it. | 2 | 2 |
| SUS Scale | 83.57 | 86.80 |
| <sup>1</sup>Note that the SUS uses a Lickert Scale with 1 equal to “Strongly Disagree” and 5 equal to “Strongly Agree”. For scoring details please see Brooke [20] |  |  |
| Letter grade and adjective rating (see Supplementary Table S1) | A/Excellent | A/Excellent |

### Emergent Themes

Several themes emerged from the analysis of the semi-structured interviews (**Table S5**) and are described below.

#### Fall risk assessment is a team effort

Providers assess fall risks both individually and as a team. Team processes involve multiple inputs throughout the patient visit, which are ultimately directed to the provider for interpretation and decision-making. In practice, participants commented on the overall benefits and potential modifications of the current algorithms and assessment techniques.

#### Hesitation about using the QuPI with certain types of patients

Participants were hesitant to use the QuPI with certain types of patients, including those using assistive devices (cane, walker, wheelchair) or with a diagnosis of diabetes, obesity, or peripheral neuropathy. There were also minor hesitations about using the QuPI tool with patients with big feet, patients that could not place their feet close together, and some patients 65 years of age or older (**Table 4**).

**Table 4.** Participants’ experience in initial use of the QuPI.

|  |  |
| --- | --- |
| Implications | 78% (14/18) liked how well the QuPI worked in terms of their clinical process. |
|  | 100% (18/18) said that, with some hesitation or qualifications for certain patients, they would use the QuPI with all patients. Reasons for hesitation included: patient age, obesity, osteoporosis, poor results on observation test, large feet, visual impairment, use of wheelchair or walker, workflow bottleneck, complaint about a recent fall or concerns, patients coming for a follow-up appointment. |
|  | 89% (16/18) agreed that the QuPI would be easily added to the vital sign assessment in their clinical practice. |
| Concerns/Discussion | 61% (11/18) stated that time constraints, such as taking off shoes, cost of the device, amount of time training staff, and patients with assistive devices would prevent the use of the QuPI. |
|  | 100% (18/18) had some hesitation about using the QuPI with certain types of patients. The concerns were related to patients over 65 years of age and patients with osteoporosis, assistive devices (wheelchair, cane, etc.), obesity, or foot issues (size, neuropathy). |
| Replacement of current scales with the new scale | 100% (18/18) stated they would like to use the QuPI in place of a scale.<br>The preferred physical location for the QuPI would be in the triage room (1 participant), vital station (15 participants), or exam room (2 participants). |
| Recommendations | Features the participants liked were ease of use, simple instructions, appropriate height and size, dual-purpose for balance and weight measurements, audible tone to indicate the end of the test, ability to |
|  | give feedback during the test, portability, and the 30-second time for the test. |
|  | Participants suggested improvements including making the scale larger, adding rails or handlebars near the tool, allowing the feet to be further apart, and having the tool flatter to the ground. |
|  | 64% (9/14) liked having the test only take 30 seconds instead of 60 seconds. The other four participants did not see a prototype with a 60-second test duration. |
|  | 67%, (12/18) mentioned other desired features for the QuPI, including automatic data entry into electronic health records; inclusion of a level to facilitate placement of the tool; addition of height measurement to the tool; color codes on the display to reflect normal, moderate, and high-risk values for easier interpretation; and placement of an outline of the feet similar to those on airport scanners. |

> *"…I might not say all my patients, but you know my hardest patients [including] all of my Medicare patients…Maybe I would just use it on 65 and older and anyone who might be just complex.”*
>
> *“I mean, I think I could see it being used on every patient versus like if we could identify a higher risk population…”*

#### Knowledge of national algorithms

Two-thirds (12/18) of the participants knew or had heard of one or more of the nationally promoted algorithms from the Centers for Disease Control and Prevention (CDC), Johns Hopkins, or the AGS [23–25]. These algorithms for fall risk assessments each begin with three similar key questions: Has the patient fallen in the past year? Does the patient feel unsteady? Does the patient have difficulty walking? If the patient does not answer ‘yes’ to any of these questions, the fall risk assessment is concluded. On the other hand, if the patient answers ‘yes’ to any of these key questions, then providers ask more in-depth questions regarding medication history and any injury from previous falls. The algorithms then proceed from this qualitative assessment to a quantitative analysis using functional mobility testing.

In terms of fall risk assessments, 50% (9/18) of the participants asked patients specifically about fall risks, and 50% (9/18) assessed patients’ gait by watching them walk from the waiting room to the examination room. Only 43% (6/14) of the participants that took vital signs for every patient at every visit said that information about fall risks or balance problems was brought up by patients. The other 57% of participants said information about fall risks was brought up explicitly by a medical assistant, nurse practitioner, or physician through direct questioning.

#### Desire to use the QuPI in place of a scale

All the participants said they would like to use the QuPI in place of a conventional weight scale. The physical location in which the participants said they would replace the current scale with the QuPI was the triage room (1 participant), the vitals station (15 participants), and the exam room (12 participants) (**Table S6**).

> *"I think in terms of workflow it would be pretty easy minus the added step. But I mean, like I said, we’ve got a patient on a scale every time they come and get their vitals. So, in terms of the workflow, no matter which clinic you’re in, they’re probably going to go into a vitals room or hallway and get their weights anyways. So just adding to that, I think it’s reasonable."*
>
> *“Since it takes the place of a scale, it’s already getting your weight so that’s nice. And then it gives you another objective, almost like a vital sign or data point for the issue of falls."*

#### The QuPI was easy to use

The participants liked that the QuPI was easy to use, provided objective data, had simple instructions, was the right height and size, has an interface that was easy to interpret and use, was dual-purpose (weight and balance), provided an audible ‘ding’ at end of the test, was portable, and took only 30 seconds to use. They also appreciated being able to give patients feedback regarding the time remaining on the test and the patients’ cooperation with the test procedure (**Table S7** and **Table S8**).

> *"It’s helpful, it gives some more objective data that the patient and me could both visually look at and see that progress was made. And it’s quick, I think that’s helpful. Easy to use." "I think it’d be easy. There’s already a weight machine, so it’s much different than them having to stand on another machine to get the weight, and then it’s small. It fits well. It’s short duration overall."*

## Discussion

The central premises underlying this work are that an inexpensive, easy-to-use QuPI system can detect significant balance deficits below the threshold of patients’ and providers’ perceptions and generate quantitative data for evidence-based assessment. Previous studies demonstrated that clinical research coordinators or other medical assistants were capable of using similar QuPI systems to detect balance changes in patients on chemotherapy that preceded patient-reported or provider-reported neurological, sensory, or motor changes [13–14]. This suggests that by overcoming barriers to QuPI implementation in clinical settings, providers can screen for balance changes more sensitively and objectively than they can using current approaches. The participants unanimously expressed acceptance of the second prototype and said they would replace their current weight scale with the modified QuPI, which suggests that the changes made between the first and second rounds of interviews made progress in overcoming barriers to implementation. In particular, several participants indicated the QuPI overcame some limitations of national algorithms, including dependence on patient recall and awareness, potential over-reliance on medications or injurious falls as evidence of poor balance, and the significant time and expertise needed to conduct functional balance testing.

> *“I do, I guess I do have concerns about [the JHFRAT]. Because, I don’t have it in front of me, it’s medicine and it’s age, and I think some people score higher than I would expect them to have a problem and I sometimes don’t think, because of the medicines, the medicines are very high I think on the list. Which does make sense, but I… Yeah, yeah in some ways I don’t [inaudible] that, yeah. It’s not perfect, at all.”*

The processes for fall risk assessments varied across the clinics in our study, despite the existence of national guidelines designed specifically to make fall risk assessments more consistent. Use of the QuPI, which can be easily integrated into existing workflows, would facilitate consistency of the process for fall risk assessments. By providing an interpretable score, the QuPI can also facilitate communication among members of the care team. In many cases, fall risk assessments are done via a team process, whereby several members of the care team each collect some information that they then share with the provider of record to use in a treatment plan. This approach has several advantages but also several disadvantages. One major advantage of the team-based approach is that it provides multiple opportunities for the provider and support staff to observe and collect information on the patient, rather than depending solely on the limited time that the provider spends with the patient. For example, medical assistants often act as an ‘extra set of eyes’ by observing the patient walk from the waiting room to the vitals station and the exam room. Observation or questioning of the patient outside the formal encounter with the provider also has the potential benefit of avoiding bias due to the Hawthorne effect, whereby the patient moves differently when they know they are being evaluated.[26] A primary disadvantage of the team approach may be a lack of consistent training for all members of the care team on how to diagnose fall risks. For the 86% (16/18) of participants who had not received formal training to work with geriatric patients (e.g., during a geriatrics fellowship), all of their training presumably came ‘on the job’ as directed by a mentor, preceptor, or provider (in the case of medical assistants or physician extenders). Inconsistency in such informal training might lead to inconsistent diagnoses of fall risk and potentially injurious falls. Use of the QuPI could potentially enhance the advantages of a team-based approach by providing a quantitative assessment that can be communicated in place of or in addition to subjective observations. Moreover, use of the QuPI would help overcome the disadvantages of inconsistent training by providing a simple-to-use tool that everyone on the care team can be trained to use reliably.

## Conclusions

The majority of participants reported that they would use the QuPI for quantitative postural control measurement with all their patients. Furthermore, all of the participants stated that they would replace their current weight scale with the QuPI. The QuPI provides a new method to quantify fall risks that was well accepted by users and was feasible to use in a clinical environment without disrupting existing workflows. The QuPI holds promise to enhance advantages and overcome barriers related to documentation, team-based workflow, and variable methods for fall risk assessments in clinical settings.

### Protection of Human and Animal Subjects

This study was performed in compliance with the World Medical Association Declaration of Helsinki on Ethical Principles for Medical Research Involving Human Subjects and was reviewed and approved by The Ohio State University Biomedical Sciences Institutional Review Board. At the time of the interview, the oral consent script was read by the interviewer to the participant, and the interviewer answered any questions from the participant. Once the participant agreed to continue, the interview began. Written consent was waived by the IRB for this study.

## Data Availability

Full transcriptions of the interviews analyzed in this study cannot be shared publicly to protect the privacy and anonymity of participants. If you wish to obtain access to this data, please contact the corresponding author.

## Author Contributions

All authors contributed to the conception of the work, interviews of participants, data analysis, and drafting of the paper.

## Acknowledgments

We thank the participants for their valuable insights.

## Supporting information captions

**S1 Table**. SUS quartile ranges and scores.

**S2 Table**. Barriers to the use of guidelines based on the Cabana Framework.

**S3 Table**. Participant descriptions of barriers to use of guidelines based on the Cabana Framework.

**S4 Table**. Recommended tool changes and modifications from the first round of interviews.

**S5 Table**. Emerging themes with supporting snippets.

**S6 Table**. Work processes associated with obtaining vital signs. **S7 Table**. Example snippets from the first round of interviews.

**S8 Table**. Example snippets from the second round of interviews.

## Notes

### Competing Interest Statement

The authors have declared no competing interest.

### Author Declarations

This study was performed in compliance with the World Medical Association Declaration of Helsinki on Ethical Principles for Medical Research Involving Human Subjects and was reviewed and approved by The Ohio State University Biomedical Sciences Institutional Review Board (study Number: 2018H0447) . At the time of the interview, the oral consent script was read by the interviewer to the participant, and the interviewer answered any questions from the participant. Once the participant agreed to continue, the interview began. Written consent was waived by the IRB for this study.

